# Regional and national estimates of children affected by all-cause and COVID-19-associated orphanhood and caregiver death in Brazil, by age and family circumstance

**DOI:** 10.1101/2025.01.31.25321479

**Authors:** Nicholas Steyn, H Juliette T Unwin, Jamie Ponmattam, Andrés Villaveces, Luiza Martins, Lorraine Sherr, Alexandra Blenkinsop, Elizaveta Semenova, Alice Stuart-Brown, André Victor Ribeiro Amaral, Oliver Ratmann, Ricardo Parolin Schnekenberg, Lucie Cluver, Susan Hillis, Laura Rawlings, Lorena Barberia, Andrea Santos Souza, Marcia C Castro, Seth Flaxman

## Abstract

Children affected by orphanhood of any cause may benefit from assessment and referral to appropriate services. Timely and accurate data can guide policy. We leveraged new data sources expanding previous reports on national COVID-19-associated orphanhood to estimate national and sub-national numbers of children newly affected by death of parents and co-residing elderly caregivers due to all-causes and to COVID-19-associated causes in 2020-2021 in Brazil.

We estimated that 1,300,000 (95% uncertainty interval 1,190,000, 1,430,000) children in Brazil experienced loss of one or multiple parents and/or co-residing caregivers 60+. 673,000 (652,000, 690,000) children were estimated to have lost one or both parents, of which 149,000 (144,000, 154,000) were COVID-19-associated; 635,000 (534,000, 758,000) children were estimated to have lost a co-residing grandparent or other kin, of which 135,000 (85,900, 199,000) were COVID-19-associated.

Orphanhood estimates varied across states. The highest all-cause rate of parental orphanhood was in Roraima, at 17.5 (95% uncertainty interval 15.6, 20.6) per 1000 children, and the lowest was in Santa Catarina, at 9.5 (8.7, 10.4) per 1000 children. COVID-19-associated orphanhood was also unevenly distributed, with Mato Grosso experiencing the greatest rate, at 4.4 (3.9, 5.3) per 1000 children, while Pará experienced the lowest rate of 1.4 (1.2, 1.8) per 1000 children.

We compared our estimates with administrative data for COVID-19-associated orphanhood (from Brazil’s civil registry offices and manually reviewed death certificates in Campinas) and found that a similar demographic distribution of orphanhood. However, our estimates suggested that administrative sources undercount orphanhood, suggesting that approximately 32% and 56% of total orphanhood was captured in the two datasets, respectively.

Our findings highlight the extent of orphanhood in Brazil and the large inequalities between states. Our comparisons with administrative data both validate our model and suggest that strengthening vital registration systems can put children at the center of public health responses globally.

## Introduction

Orphanhood (death of one or both parents) and caregiver death among children aged 0-17 years, across causes of parent and caregiver death, can have severe immediate and life-long consequences for children. Over 15 years ago, WHO classified orphanhood as an adverse childhood experience (ACE), linked to long-term mental health risks (Kessler et al. 2010). As an ACE, parent or caregiver loss may have lifelong consequences, including increased risks of suicide, post-traumatic stress disorder, violence, insecure housing, institutionalization, poverty, and chronic and infectious diseases (Kidman and Palermo 2016; Villaveces et al. 2024). Children experiencing orphanhood (“orphans”) are defined as those aged 0-17 whose father or mother died during childhood or adolescence, while the term “double orphan” refers to children who have lost both parents (Sherr et al. 2008). We also consider the loss of co-residing elderly kin (defined as individuals aged 60+ living in the same household), who often play a critical role in childcare and support (Buchanan and Rotkirch 2018; Hillis, Unwin, Chen, et al. 2021).

Orphanhood from any cause can dramatically affect child wellbeing. Therefore, it is helpful to understand the number of children bereft of parents or caregivers due to COVID-19 within the context of all children whose parents and caregivers die. Not only do the children face the loss of a caregiver, but they are also potentially affected by a myriad of additional stressors. Surviving parents and caregivers of orphans may be deeply bereaved by the death and their child-caregiver care and child interactions may be negatively affected. The circumstances surrounding the death may well impact the child. The death itself, and at times the illness and circumstances preceding the death, can have secondary ramifications such as economic losses due to reduced income, cost associated with alternate care arrangements, and increased health care expenditure. Children may experience multiple changes not only in their caregiving, but in their residence if moved to alternative carer domiciles or institutions. They also may experience changes in their family composition, especially if siblings are separated and readjustments to new family structures and conditions are required (Goldman et al. 2020).

A variety of interventions to improve the lives of orphans and vulnerable children have been studied (Thomas et al. 2020), with promising evidence for social protection combined with parenting support for surviving caregivers (Hillis, Unwin, Cluver, et al. 2021). Delivering protection and support for children orphaned from any cause is only possible if bereaved children can be identified and linked to care in a timely manner. Brazil is unique in the world in requiring a field on death certificates recording child dependents in the household.

In Brazil, this simple death certificate data holds out the possibility of swiftly identifying children affected by orphanhood and caregiver death, so that their health, social, and economic needs can be assessed and those in need can be linked to health, psychosocial, parenting, and economic services (Flaxman et al. 2023). Andrea Santos Souza (a public prosecutor and co-author), aware of the scale of COVID-19-associated orphanhood in Brazil (Hillis, Unwin, Chen, et al. 2021), obtained death certificates to systematically identify and link COVID-19 orphans to economic support in Campinas (a municipality in São Paulo State). Administrative data on orphanhood is rare globally, so its existence in Brazil is a unique opportunity both to provide support to orphaned children and to validate statistical models for estimating orphanhood. Furthermore, marked subnational disparities in COVID-19-associated cases and deaths in Brazil (Castro et al. 2021) have been linked to differing public health interventions, levels of preparedness (Rocha et al. 2021) and pre-existing inequities (Brizzi et al. 2022). The degree to which all-cause orphanhood and the subset of COVID-associated orphanhood distributions also reflect these subnational public health disparities has not been described.

In spite of its acute and enduring lifelong consequences, the threat of all-cause orphanhood has been largely invisible as a major global public health priority. However, orphanhood is a recognized threat to children in the context of disease epidemics, including HIV/AIDS (Cluver et al. 2023; Belsey and Sherr 2011), COVID-19 (Hillis, Unwin, Chen, et al. 2021) and Ebola (Evans and Popova 2015). In Brazil, similarly, although invisibility characterizes all-cause orphanhood at federal, state, and municipal levels, attention to policy reform has accelerated for COVID-19-, emergency-, and disaster-linked orphanhood. Sparked by the large-scale repercussions of the COVID-19 health emergency on families, the Ministry of Social Development announced a national program for orphans of health emergencies and natural disasters, targeted for launch in 2024. Legislation is also under discussion in the Brazilian Senate and Congress to provide special pensions to emergency-linked orphanhood and ensure support for social assistance programs guaranteeing access to educational, psychological and health services (“PL 2291/2021 - Senado Federal” 2023; “Portal da Câmara dos Deputados” 2021a; “Portal da Câmara dos Deputados” 2021b; “Portal da Câmara dos Deputados” 2023). At the sub-national level, a consortium of states in the Northeast (Maxmeio 2021), and municipalities, such as Campinas, passed legislation to identify, assess, and link COVID-19 orphans, to grief counseling, economic, educational, and psychosocial support (“Lei Ordinária 16200 2022 de Campinas SP” 2022). Such policy commitments to emergency-linked orphanhood in Brazil lay a strong foundation for expanding eligibility to include coverage of children with acute needs who are orphaned for any cause and sets a precedent for future crisis and emergency responses.

Given the enduring impact of all-cause orphanhood on children and the context of escalating polycrisis threats to children and families, it is helpful to understand the nature and extent of the burden, geographical and regional variances, and the drivers not only of orphanhood but also of co-residing caregiving loss among children and the context in which the caregiver(s) are lost, thus facilitating better provisioning of support and care for affected children (Hillis, Unwin, Chen, et al. 2021). Therefore, we leveraged new data sources to estimate national and sub-national numbers and rates of children newly affected by death of parents and co-residing grandparents or older kin due to both all-cause orphanhood and to COVID-19-associated orphanhood in 2020-2021 in Brazil. We aim for our findings to inform immediate and long-term policy reform and ensure children are seen and cared for as part of future emergency and pandemic crisis preparation.

## Materials and Methods

We built on existing methods (Hillis, Unwin, Chen, et al. 2021) to estimate all-cause and COVID-19-associated orphanhood and caregiver death in children aged 0 to 17 in Brazil at the state and national level for 2020 and 2021. Our calculations rely on estimates of fertility rates, household composition, and excess mortality in adults. We additionally considered the impact of deaths of co-residing individuals aged 60 years and older (60+) as a proxy for the loss of custodial grandparents and other older kin. All our estimates are structured by age-group (in 10-year groups limited by the disaggregation of the all-cause mortality data) and sex, with further disaggregation by month, state (26 federative units and 1 federal district), and current age-of-child where necessary. The Pesquisa Nacional de Saúde (PNS - National Health Survey, 2019) was used to estimate fertility rates for men aged 18+ and the household composition of adults 60+. Live births and population data were used to produce estimates of female fertility (IBGE 2022; “Sistema de Informação Sobre Nascidos Vivos – Sinasc - OPENDATASUS” 2023). All-cause mortality estimates for Brazil for 2020-2021 are available in Ponmattam et al. (2024).

The number of children experiencing the death of a mother and/or a father was estimated as the number of children currently aged 0 to 17 who were born to parents who died, assuming that age-structured fertility rates are independent of the propensity for a given parent to die. The estimate was performed at the state, parent age-group, parent sex, and year-of-death level, and additionally at the age-of-child or month-of-death level where necessary, before being aggregated. Estimates of children who experienced grandparent or older kin loss were created by summing the number of children living in households where a single person aged 60+ died, where multiple people aged 60+ died, and where someone aged 60+ died and where no adults aged 18-59 were living in the household (indicating the loss of a direct caregiver).

Uncertainty in live-births reporting, survey data used to estimate male fertility and older persons household composition, and mortality was accounted for via bootstrapping (Unwin et al. 2022). Individual-level uncertainty in female fertility estimates was accounted for by assuming Poisson noise about the population-level bootstrap samples, while individual-level uncertainty in male fertility and household composition was derived directly from the survey data. The mean and the 2.5^th^ and 97.5^th^ quantiles of the bootstrap samples provided our central estimates and uncertainty intervals. Full details are provided in supplementary material section 2.

### All-cause and excess mortality

Expected all-cause mortality was predicted using a Poisson generalized linear mixed model that was trained on 2013-2019 historical mortality data obtained from Brazil’s Mortality Information System (Ponmattam et al. 2024). The fixed parameters were 1-year lagged all-cause mortality rates, and month and age group indicators; states were included as random parameters so that each state had a random intercept and slope on the year variable. Each sex was modeled separately. Excess mortality is estimated as the number of observed deaths which are in excess of the expected number for a given time and place (Ponmattam et al. 2024). Our time period is one in which COVID-19 was a major cause of death, likely accounting for the vast majority of excess mortality (Ponmattam et al. 2024). Excess mortality is a more accurate representation of the overall impact of the COVID-19 pandemic than COVID-19-reported deaths (Msemburi et al. 2023), which were under-reported during the ongoing epidemic when health systems were overloaded (Brizzi et al. 2022), thus enabling us to come closer to the true magnitude of children whose parents or caregivers died during the COVID-19 pandemic. Estimates of all-cause and excess mortality by month, age-group, sex, and state (and federal district) were obtained for the period from January 2020 through December 2021 (Ponmattam et al. 2024). We report estimates associated with excess deaths as “COVID-19-associated orphanhood”, while estimates associated with all-cause mortality are reported as “all-cause orphanhood”.

Confidence intervals for monthly data were not available, and as such we did not further account for uncertainty in monthly excess mortality estimates in our analyses. Monthly data were only used when presenting trends in orphanhood results by month in 2020 and 2021.

### Estimating female fertility

Female cumulative child fertility rates were estimated by summing the relevant annual age-specific fertility rates for females currently aged 15 or above. A weighted average by population size was used to aggregate these estimates from single-year age-groups to 10-year age-groups. This aggregation was also performed at the month and state-level where necessary. To determine the number of children currently aged 0 to 17 years born to women (female cumulative child fertility rate) we obtained live births data from 2003 through 2020 (“Sistema de Informação Sobre Nascidos Vivos – Sinasc - OPENDATASUS” 2023). These were used to estimate annual fertility rates by dividing the total number of births recorded to women aged 10 through 65 by individual age in each year and state by the corresponding population sizes (IBGE 2018). Historical population sizes were only available in 5-year age-groups so we used the Sprague interpolation method (De Kerf 1975) to expand these into single ages where necessary. Cumulative child fertility rates were then calculated by summing the relevant age-specific fertility rates for females currently aged 15 or above.

Various factors result in live births data slightly undercounting the actual number of births (“Sistema de Informação Sobre Nascidos Vivos – Sinasc - OPENDATASUS” 2023). Therefore, we adjusted our totals using under-reporting estimates (IBGE 2022). The under-reporting estimates range from 0.52% (in mothers aged 38) to 5.32% (in mothers aged 49). Under-reporting estimates were provided for 2020 only and without associated uncertainty. To capture the potentially high level of uncertainty, we assumed that true age-structured under-reporting of live births could range from 0 through 2 times the estimated level. We also account for child mortality. See supplementary material section 2.2 for further details.

### Estimating male fertility

Age and sex disaggregated live birth data were not available for men before 2010, so the 2019 PNS health survey was used to estimate male fertility. Here, we adapted the commonly used own child method (Timæus 2021) based on responses to the PNS 2019 household survey (“Sistema de Informação Sobre Nascidos Vivos – Sinasc - OPENDATASUS” 2023).

Briefly, if the primary respondent (a single randomly selected individual aged 15+ in each household) to PNS 2019 was male (N=44,752), they were asked the number of children they had, the age of their youngest living child, and the year of birth of their oldest child. 30,579 (68.3%) eligible men reported having at least one child. When a father reported having three or more children with the youngest < 18 years old and the oldest child ≥ 18 years-old (N=3,408, 8.0% of eligible men), the total number of children 0 to 17 years old remained uncertain. For these observations, we assigned random ages to the middle child or children uniformly between the age of the youngest and the age of the oldest child. To quantify uncertainty in male fertility, we impute middle-child ages for 1,000 possible PNS surveys, and for each possible survey, we generate a sample of possible numbers of children for fathers in each age-group and state combination. We also account for child mortality. Further details are in supplementary material section 2.3.

From 2010 onwards, the age of the father was included on some live birth records. We leverage these data in supplementary section 3.3 to provide validation to our survey-derived estimates.

After 2010, our estimates were validated by comparing to estimates from live birth data (see supplementary material section 2.3).

### Single Orphanhood

Samples of orphanhood resulting from the death of a parent in a specific age-group, sex, state, and year are constructed by first sampling the number of deaths (excess or all-cause) that occurred in adults in this group. Then, for each parent that died, we sample (from estimates of female or male cumulative child fertility rates) the number of children aged 0 to 17 that they left behind. Totaling all children left behind (in each age-group, sex of parent, state, and year combination) produces a single sample from the distribution of orphanhood for that combination.

Repeating this procedure produces a collection of samples of orphanhood that approximate the distribution of orphanhood. Our central estimates are the average of these samples, while the 2.5th and 97.5th quantiles define our 95% uncertainty intervals (U.I.). Aggregated estimates of orphanhood (for example, total orphanhood, or orphanhood by state) were calculated by summing over the relevant samples and then calculating the average and quantile values.

### Double Orphanhood

To account for double orphanhood (the loss of both parents) we estimated the number of male-female couples where both have died. We do not explicitly account for adult mortality pre-2020, so our definition of double orphanhood includes pre-existing single orphans who became double orphans in 2020 or 2021.

We first estimated the age-distribution of male-partners of women in each age-group using PNS survey data, assuming that the age-distribution of within-household male-partners was similar to the age-distribution of out-of-household male partners. While this assumption may not hold in the real setting, we do not expect it to have a meaningful impact on our results given the overall low rates of double orphanhood.

The double excess mortality for women in the i^th^ age-group with male partners in the j^th^ age-group was estimated as follows: for each excess death in a woman, sampling an age-of-partner j from the distribution estimated above, and assigning their partner as an excess death with probability Ej/Nj, where Ej is the total number of excess deaths in men in age-group j, and Nj is the total number of men in age-group j. Repeating this procedure and taking the average produced our central estimates of double excess mortality. Our approach assumes independence between two events: death of a mother and death of a father. In reality, there is likely to be a positive correlation between these events; if we took this into account, it would increase our estimates of double orphanhood.

As there is insufficient information to estimate joint fertility rates, we used the estimated fertility rate associated with the female partner, because it comes from registry data and so is more reliable than our male fertility rate estimates. Accurate estimates for female fertility are important because of the more restricted window of female fertility. A limitation of this strategy is that it ignores the fact that a mother in a given age-group is less likely to have children aged 0 to 17 if their partner is older, which is also when both parents are more likely to have died. If we took this into account, it would decrease our estimates of double orphanhood.

The limitations listed above will have a small impact on total orphanhood estimates because double orphanhood is very small relative to single orphanhood. Full details are given in the supplementary material.

### Estimating the loss of co-residing grandparent or older kin

We used the PNS 2019 survey to consider household-based estimates of orphanhood, allowing us to estimate the number of children that lost a co-residing grandparent or older kin. We used mortality estimates to assign excess or all-cause deaths to individuals aged 60+ that featured in the survey with probability E/N, where E is the number of excess deaths that occurred in individuals in a specific age-group, sex, state, and year, and N is the corresponding population size.

We then used the R *survey* package (“Survey: Analysis of Complex Survey Samples” 2023) to estimate the total number of children living in households where:

- At least one elderly co-residing person has died (“any”)
- A single elderly co-residing person has died (“single”)
- Multiple elderly co-residing people have died (“multiple”)
- At least one elderly co-residing person has died and there are no adults aged 18-59 (“direct”, representing the loss of a direct caregiver)

The *svytotal* function from the *survey* package in R was used to estimate a survey-weighted mean and standard error for total orphanhood due to the loss of co-residing elderly. This process was repeated 100 times, at each iteration reassigning excess deaths, estimating a new mean and standard error, and taking 10 samples of total orphanhood from the approximately Normal distribution (with the corresponding estimated mean and standard error). This process produced 1000 samples of orphanhood due to the loss of co-residing elderly that accounted for uncertainty in excess mortality and household composition.

To estimate the total number of children experiencing either parental orphanhood or the loss of co-residing grandparents or older kin, we add the estimated number of children experiencing parental orphanhood and the estimated number of children that lost co-residing grandparents or older kin, and subtract the small number of children that experienced both, estimated using unweighted PNS data by state (supplementary section 2.6).

### Monthly orphanhood

Uncertainty estimates were unavailable for excess mortality by month, restricting our ability to explicitly account for double orphanhood. When presenting estimates by month we do so without associated uncertainty and scaled such that the total matches the expected total from estimates calculated from our full methods.

### Administrative Data

We obtained two sources of administrative data to situate our estimates. Arpen-Brasil, which represents Brazil’s Civil Registry offices, used administrative records to link birth and death certificates in 2021 for children aged 6 or under (“Covid já deixou órfãs ao menos 12 mil crianças de até 6 anos, indicam cartórios” 2021) and shared summary tables of their results. This dataset, released in October 2021, consists of children who experienced orphanhood between 18 March 2020 and 24 September 2021. A total of 12,566 orphans (including maternal, paternal, and double) were identified in this period for children aged 6 or under: n = 12,211 for children whose parents’ death have been recorded as COVID-19 and n = 355 for Severe Acute Respiratory Syndrome (SRAG in Portuguese) deaths. While COVID-19 is a likely cause of death for the SRAG deaths, we restrict our analysis to the COVID-19-confirmed deaths.

Co-author Andrea Santos Souza and a team of research assistants manually reviewed all death certificates in 2020 and 2021 listing COVID-19 as the cause of death in Campinas and created a list of affected children 0-17. They uncovered a total of 481 children aged 0-17 listed as child dependents in the household of adults who had died from COVID-19.

## Results

### All-cause orphanhood

We estimate that, from January 2020 through December 2021, a total of 1,300,000 (95% uncertainty interval 1,190,000, 1,430,000) children in Brazil experienced loss of one or multiple parents and/or grandparent or older kin (two-year incidence). Of these, 673,000 (95% uncertainty interval (U.I.) 652,000 - 690,000) children lost one or both parents due to any cause of death (Table 1, Figure 1). Consistent with the global mortality impacts of COVID-19 which affected men disproportionately, and the tendency for men to have children at older ages, paternal orphanhood (496,000 children, or approximately 73.8% of all orphaned children) was estimated to be more frequent than maternal orphanhood (174,000 children, or approximately 25.8% of all orphaned children). The remaining 2,730 children (0.4%) were orphaned due to the loss of both parents. Orphanhood was estimated to be greater in older children than younger children, particularly for the loss of mothers and the loss of both parents (Figure 2).

**Figure 1.**
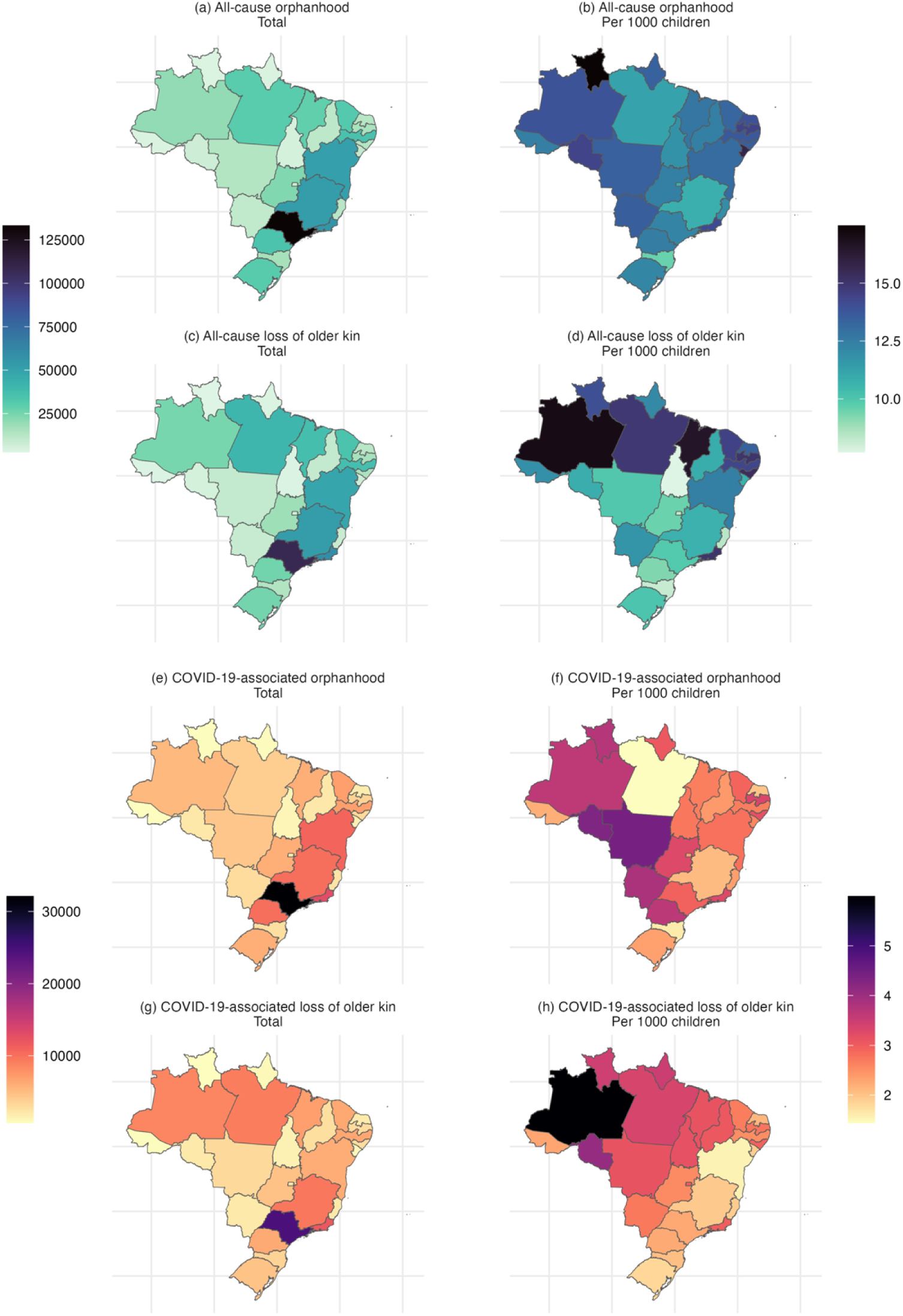
Estimates of (a, b) the number of children experiencing orphanhood due to all causes, (c, d) the number of children that lost a co-residing grandparent or other older kin due to all causes, (e, f) the number of children experiencing orphanhood due to COVID-19-associated causes, and (g, h) the number of children that lost a co-residing grandparent or other older kin due to COVID-19-associated causes. Left-hand figures show absolute estimates, right-hand figures show estimates per 1000 children in the given state.

**Figure 2.**
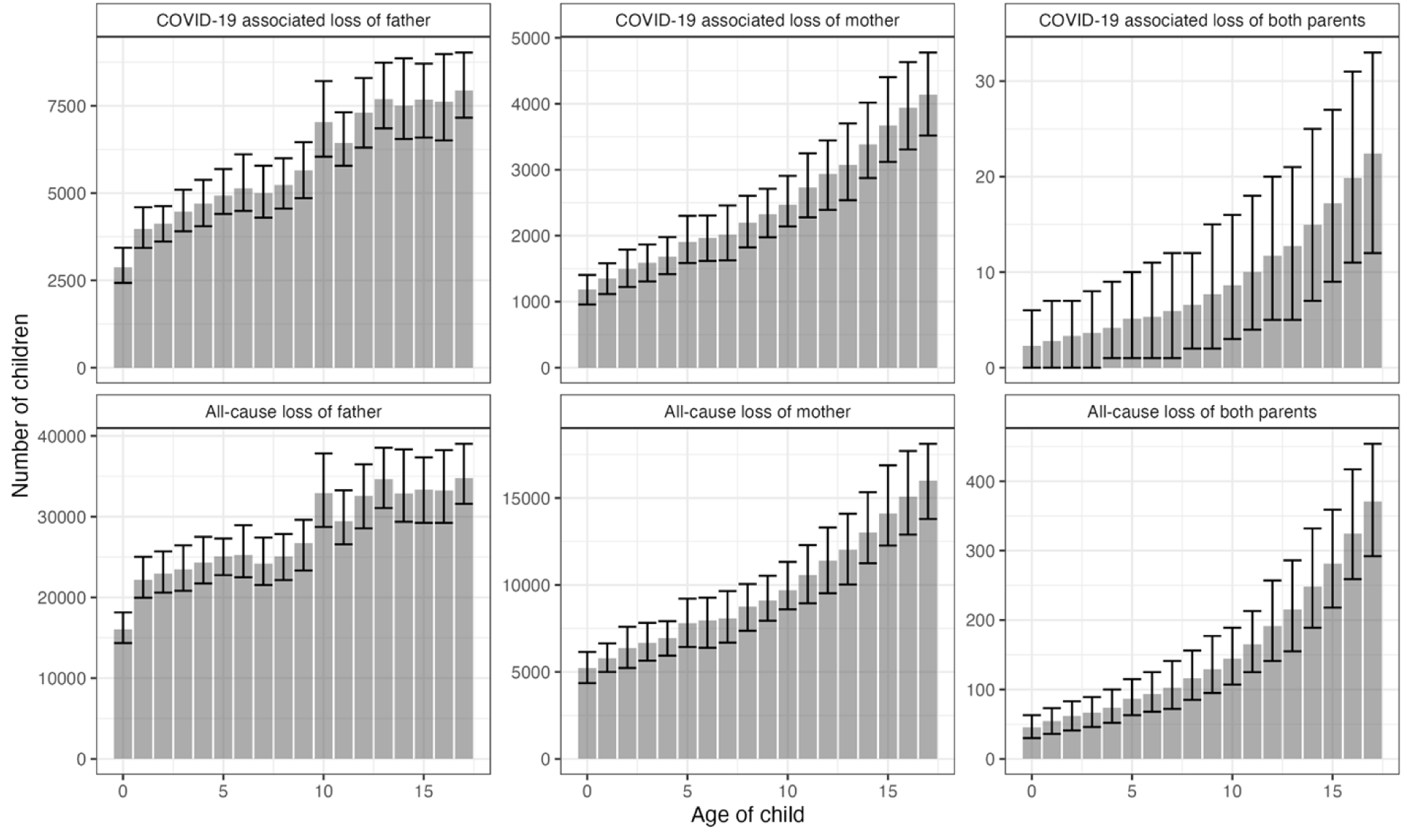
Age distribution of children experiencing orphanhood in Brazil from January 2020 through December 2021 by family circumstance (death of father, death of mother, death of both parents). COVID-19 associated orphanhood is shown in the top panels, all-cause orphanhood is shown in the bottom panels.

**Table 1.**
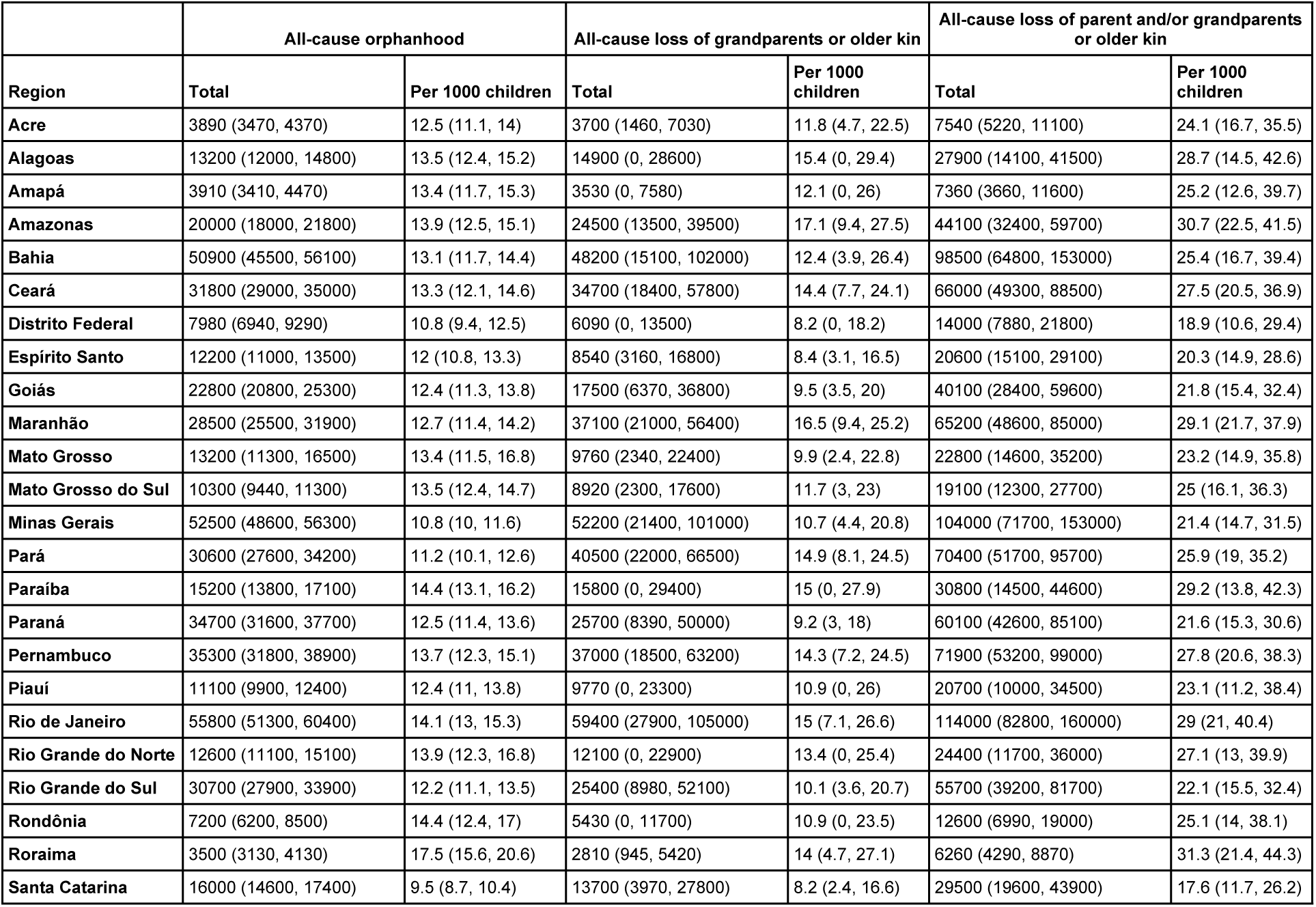
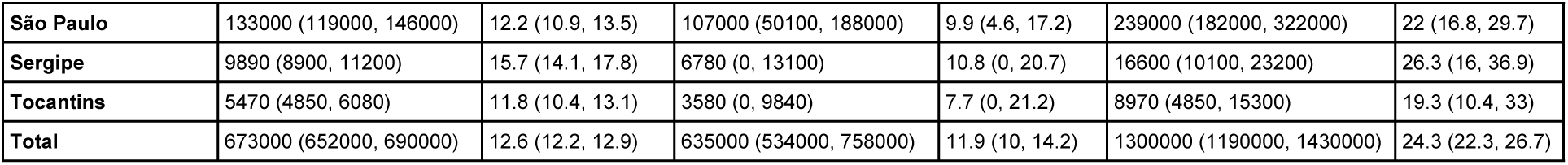
Estimates of orphanhood due to any cause of parental death and estimates of the number of children that lost a co-residing grandparent or other older kin.

In absolute terms, the three worst affected states were São Paulo with a total of 133,000 (119,000-146,000), Rio de Janeiro with a total of 55,800 (51,300-60,400), and Minas Gerais with a total of 52,500 (48,600-56,300) children experiencing all-cause orphanhood. These are the three most populous states of Brazil. When calculating state-level orphan incidence rates per 1,000 children, the three worst affected regions were the state of Roraima with 17.5 per 1,000 children (15.6-20.6), Sergipe with 15.7 per 1,000 (14.1-17.8), and Rondônia with 14.4 per 1,000 (12.4-17.0).

Over the same time period, we estimate a total of 635,000 (534,000 - 758,000) children in Brazil lost at least one co-residing grandparent or older kin (an adult living in the same household aged 60+) due to any cause of death (Table 1, Figure 1). Of these, 16,200 (3,350 - 46,600) lost multiple co-residing grandparents and/or older kin, and 91,800 (53,700 - 150,000) lost a grandparent or older kin with direct responsibility (i.e. there were no adults aged 18-59 living in the same household, although there may have been other grandparents). A relatively small number of these children may also have lost a parent – we account for this in Table 1.

In absolute terms, the three worst affected states were São Paulo with a total of 107,000 (50,100 - 188,000) children losing at least one co-residing grandparent or older kin, Rio de Janeiro with a total of 59,400 (27,900 – 105,000) children affected, and Minas Gerais with a total of 52,200 (21,400 - 101,000) affected. Incidence rates of orphanhood per-1,000-children due to death of a co-residing grandparent or older kin was worst in the state of Amazonas with 17.1 per 1,000 children 9.4, 27.5), Maranhão with 16.5 per 1,000 children (9.4 - 25.2), and Alagoas with 15.4 per 1,000 children (0, 29.4). Tables for all categories and stratifications of orphanhood can be found in supplementary section 4.

### COVID-19-associated orphanhood

We estimate that, from January 2020 through December 2021, a total of 284,000 (95% uncertainty interval 235000, 348000) children in Brazil experienced loss of one or multiple parents and/or grandparents or older kin (two-year incidence) due to COVID-19-associated causes (two-year incidence).

We estimate that from January 2020 through December 2021, a total of 149,000 (144,000 - 154,000) children in Brazil experienced COVID-19-associated orphanhood as estimated through excess mortality (Table 2, Figure 1). Of these, 105,000 (100,000 - 109,000) were orphaned due to the loss of a father accounting for 70.5% of all orphans, 43,800 (41,600 - 45,800) were orphaned due to the loss of a mother accounting for 29.4% of all orphans, and 160 (130 - 200) were orphaned due to the loss of both parents accounting for 0.1% of all orphans. Like all-cause related orphanhood, COVID-19-associated orphanhood was estimated to be greater in older children than younger children (Figure 2).

**Table 2.** Estimates of orphanhood due to COVID-19-associated deaths and estimates of the number of children that lost a co-residing grandparent or other older kin.

| Region | COVID-19-associated orphanhood |  | COVID-19-associated loss of grandparents or older kin |  | COVID-19-associated loss of parent and/or grandparents or older kin |  |
| --- | --- | --- | --- | --- | --- | --- |
|  | Total | Per 1000 children | Total | Per 1000 children | Total | Per 1000 children |
| Acre | 689 (534, 859) | 2.2 (1.7, 2.7) | 720 (0, 2690) | 2.3 (0, 8.6) | 1410 (621, 3340) | 4.5 (2, 10.7) |
| Alagoas | 3070 (2690, 3520) | 3.2 (2.8, 3.6) | 2860 (127, 10000) | 2.9 (0.1, 10.3) | 5920 (3100, 12900) | 6.1 (3.2, 13.2) |
| Amapá | 892 (719, 1080) | 3.1 (2.5, 3.7) | 1020 (0, 4190) | 3.5 (0, 14.4) | 1910 (842, 5050) | 6.5 (2.9, 17.3) |
| Amazonas | 5220 (4530, 5900) | 3.6 (3.1, 4.1) | 8620 (2770, 18300) | 6 (1.9, 12.7) | 13800 (7810, 23300) | 9.6 (5.4, 16.2) |
| Bahia | 10800 (9830, 12000) | 2.8 (2.5, 3.1) | 6030 (0, 30200) | 1.6 (0, 7.8) | 16900 (10200, 40900) | 4.3 (2.6, 10.5) |
| Ceará | 6970 (6340, 7700) | 2.9 (2.6, 3.2) | 6430 (895, 18700) | 2.7 (0.4, 7.8) | 13400 (7790, 25600) | 5.6 (3.2, 10.7) |
| Distrito Federal | 1750 (1430, 2180) | 2.4 (1.9, 2.9) | 2230 (0, 7790) | 3 (0, 10.5) | 3980 (1690, 9660) | 5.4 (2.3, 13) |
| Espírito Santo | 2390 (2080, 2740) | 2.4 (2, 2.7) | 1950 (0, 6840) | 1.9 (0, 6.7) | 4340 (2300, 9420) | 4.3 (2.3, 9.3) |
| Goiás | 6010 (5370, 6830) | 3.3 (2.9, 3.7) | 4670 (0, 18200) | 2.5 (0, 9.9) | 10700 (5840, 24200) | 5.8 (3.2, 13.2) |
| Maranhão | 5960 (5210, 6820) | 2.7 (2.3, 3) | 6980 (1580, 17500) | 3.1 (0.7, 7.8) | 12900 (7440, 23600) | 5.8 (3.3, 10.5) |
| Mato Grosso | 4350 (3780, 5230) | 4.4 (3.9, 5.3) | 3060 (0, 12900) | 3.1 (0, 13.2) | 7400 (4090, 17000) | 7.5 (4.2, 17.3) |
| Mato Grosso do Sul | 2930 (2600, 3290) | 3.8 (3.4, 4.3) | 2070 (0, 7930) | 2.7 (0, 10.4) | 4990 (2870, 10700) | 6.5 (3.8, 14) |
| Minas Gerais | 10200 (9210, 11100) | 2.1 (1.9, 2.3) | 9510 (60, 36300) | 2 (0, 7.4) | 19700 (10400, 46300) | 4 (2.1, 9.5) |
| Pará | 3930 (3210, 4820) | 1.4 (1.2, 1.8) | 9110 (1850, 23800) | 3.4 (0.7, 8.8) | 13000 (5660, 28700) | 4.8 (2.1, 10.5) |
| Paraíba | 3500 (3070, 3970) | 3.3 (2.9, 3.8) | 2910 (78, 10400) | 2.8 (0.1, 9.9) | 6400 (3490, 13900) | 6.1 (3.3, 13.2) |
| Paraná | 10200 (9280, 11100) | 3.7 (3.3, 4) | 6260 (165, 21500) | 2.3 (0.1, 7.7) | 16400 (10300, 31700) | 5.9 (3.7, 11.4) |
| **Pernambuco** | 6880 (6100, 7710) | 2.7 (2.4, 3) | 6480 (503, 20100) | 2.5 (0.2, 7.8) | 13400 (7260, 26900) | 5.2 (2.8, 10.4) |
| **Piauí** | 2170 (1830, 2510) | 2.4 (2, 2.8) | 2730 (0, 10900) | 3 (0, 12.2) | 4900 (2190, 12800) | 5.5 (2.4, 14.3) |
| **Rio de Janeiro** | 12900 (11600, 14100) | 3.3 (2.9, 3.6) | 11800 (1030, 34700) | 3 (0.3, 8.8) | 24600 (13600, 47600) | 6.2 (3.5, 12.1) |
| **Rio Grande do Norte** | 1800 (1500, 2160) | 2 (1.7, 2.4) | 1910 (0, 7270) | 2.1 (0, 8.1) | 3710 (1740, 8990) | 4.1 (1.9, 10) |
| **Rio Grande do Sul** | 5900 (5180, 6670) | 2.3 (2.1, 2.6) | 4570 (0, 18800) | 1.8 (0, 7.5) | 10500 (5580, 24600) | 4.1 (2.2, 9.8) |
| **Rondônia** | 2160 (1800, 2630) | 4.3 (3.6, 5.3) | 2040 (201, 6700) | 4.1 (0.4, 13.4) | 4190 (2240, 8830) | 8.4 (4.5, 17.7) |
| **Roraima** | 745 (566, 924) | 3.7 (2.8, 4.6) | 692 (55, 2230) | 3.5 (0.3, 11.1) | 1430 (741, 2970) | 7.2 (3.7, 14.8) |
| **Santa Catarina** | 2720 (2400, 3110) | 1.6 (1.4, 1.9) | 3430 (0, 13300) | 2.1 (0, 7.9) | 6140 (2670, 16100) | 3.7 (1.6, 9.6) |
| **São Paulo** | 32100 (27700, 35600) | 3 (2.5, 3.3) | 24700 (3180, 68000) | 2.3 (0.3, 6.3) | 56700 (34500, 101000) | 5.2 (3.2, 9.3) |
| **Sergipe** | 1630 (1350, 1940) | 2.6 (2.1, 3.1) | 1200 (0, 5410) | 1.9 (0, 8.6) | 2830 (1520, 7180) | 4.5 (2.4, 11.4) |
| **Tocantins** | 1270 (1070, 1480) | 2.7 (2.3, 3.2) | 1460 (0, 5940) | 3.1 (0, 12.8) | 2730 (1200, 7150) | 5.9 (2.6, 15.4) |
| **Total** | 149000 (144000, 154000) | 2.8 (2.7, 2.9) | 135000 (85900, 199000) | 2.5 (1.6, 3.7) | 284000 (235000, 348000) | 5.3 (4.4, 6.5) |

In absolute terms, the three worst affected states were São Paulo with a total of 32,100 (27,700 - 35,600), Rio de Janeiro with a total of 12,900 (11,600 - 14,100), and Bahia with a total of 10,800 (9,830 - 12,000) children experiencing COVID-19 orphanhood. Excluding Minas Gerais (the second most populous state), these are the three most populous states in Brazil. However, on a per-1,000-children basis, the three worst affected states were Mato Grosso with 4.4 (3.9 - 5.3) orphans per 1,000 children, Rondônia with 4.3 (3.6 - 5.3) orphans per 1,000 children, and Mato Grosso do Sul with 3.8 (3.4 - 4.3) orphans per 1,000 children.

Figure 3 presents monthly estimates of the number of children experiencing COVID-19-associated orphanhood, suggesting the impact of the Gamma variant of COVID-19 (also called the P.1 variant) that became predominant in the population during February 2021 (Gangavarapu et al. 2023). This variant caused widespread infection in the city of Manaus, the capital and largest city of Amazonas state (Faria et al. 2021; Brizzi et al. 2022), and is likely responsible for the high peak incidence orphanhood estimated in this state in January 2021. Figure 3 also shows that the state of São Paulo (the state with the greatest number of children, 10.8m), had an orphanhood rate approximately half that of Minas Gerais (the state with the next highest number of children, 4.9m), resulting in the two states experiencing comparable levels of orphanhood in absolute terms. Only between March 2021 and July 2021 was orphanhood in absolute terms significantly greater in the state of São Paulo.

**Figure 3.**
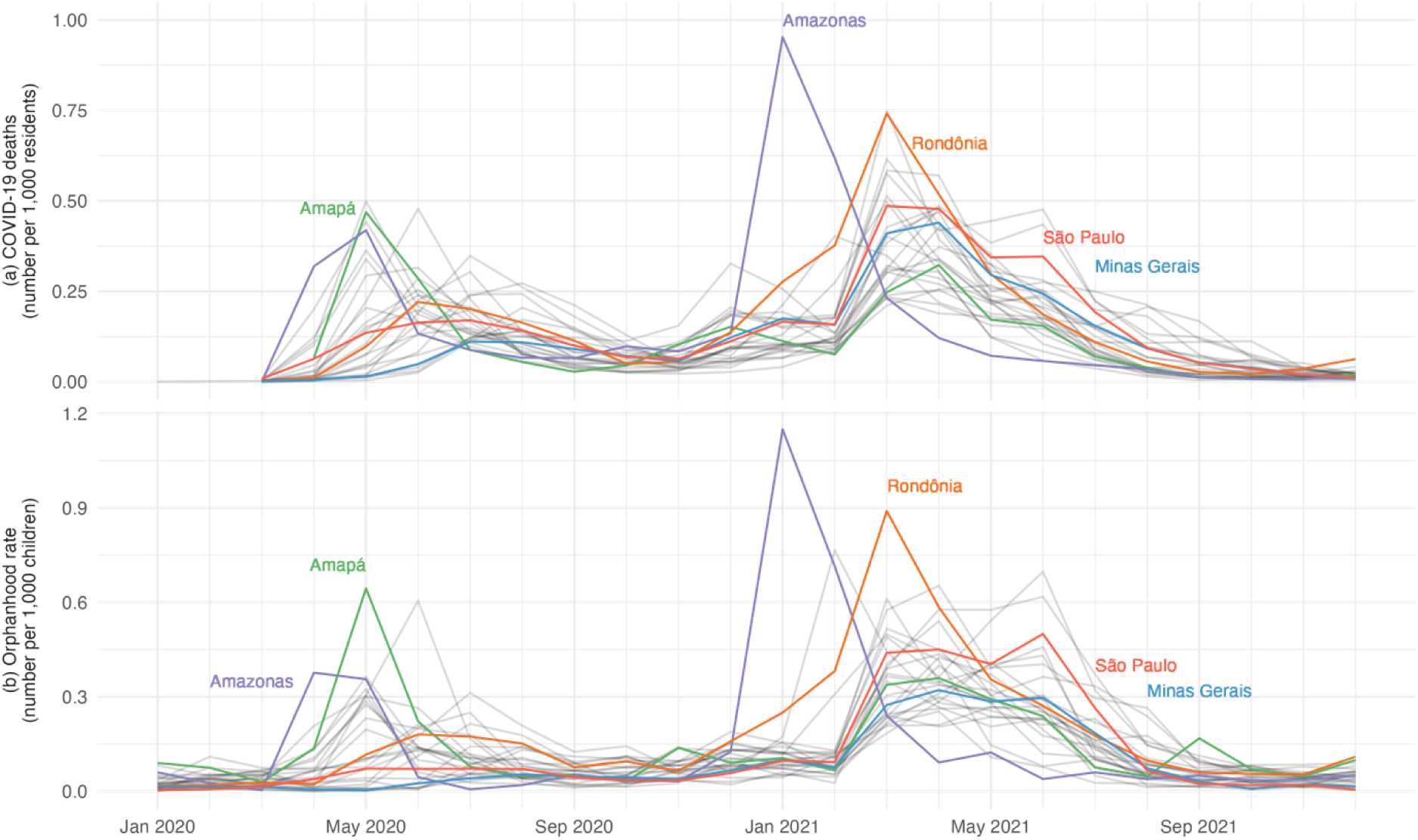
COVID-19 reported deaths (a) by state for all ages per 1,000 residents (OPENDATASUS 2022). Monthly estimates of the number of children (b) ages 0 to 17 years-old experiencing COVID-19 associated orphanhood (loss of one or both parents due to excess mortality) in Brazil from January 2020 through December 2021 per 1,000 children. Note that while there is a striking correspondence between the two sets of time series, our estimates of orphanhood in (b) were not derived from (a).

Over the same time period, we estimate a total of 135,000 (85,900 - 199,000) children in Brazil lost at least one co-residing grandparent or older kin (an adult living in the same household aged 60+) due to COVID-19-associated mortality (Table 2, Figure 1). Of these, 540 (0 - 4,130) lost multiple co-residing grandparents and/or older kin, and 21,300 (5,870 - 62,900) lost a grandparent or older kin with direct responsibility (i.e. there were no adults aged 18-59 living in the same household, although there may have been other grandparents).

In absolute terms, the three worst affected states were São Paulo with a total of 24,700 (3,180 – 68,000) children losing at least one co-residing grandparent or older kin, Rio de Janeiro with a total of 11,800 (1,030 - 34,700) children affected, and Minas Gerais with a total of 9,510 (60 - 36,300) affected. Incidence rates of orphanhood per-1,000-children due to death of a co-residing grandparent or older kin was worst in the state of Amazonas with 6.0 per 1,000 children (1.9 - 12.7), Rondônia with 4.1 per 1,000 children (0.4 - 13.4), and Amapá with 3.5 per 1,000 children (0 - 14.4).

### Comparisons with administrative data

#### Arpen-Brasil Civil Registry data

Of the 12,211 COVID-19 deaths of children aged 6 or under, the sex of the parent was recorded as male in 8,145 (66.7%) of cases and female in 3,960 (32.4%) of cases. Double orphans (loss of both parents) accounted for 103 (0.8%) children. These proportions are similar to our estimates of 70.5% paternal versus 29.4% maternal orphans for COVID-19 associated orphans. We estimated the loss of both parents to account for only 0.1% of orphaned children, a lower percentage than in the Arpen-Brasil dataset, but we note that we were unable to verify whether double orphans could have been double counted in some cases in the Arpen-Brasil dataset. The Arpen-Brasil data are reported by date-of-death from 18 March 2020 through 24 September 2021. Of these, 12,010 (98.4%) were reported from 1 April 2020 through 31 August 2021. We calculated the proportion of total orphanhood over this latter period that occurred by month for the civil registry data and our estimates (Figure 4) and found similar proportions over time.

**Figure 4.**
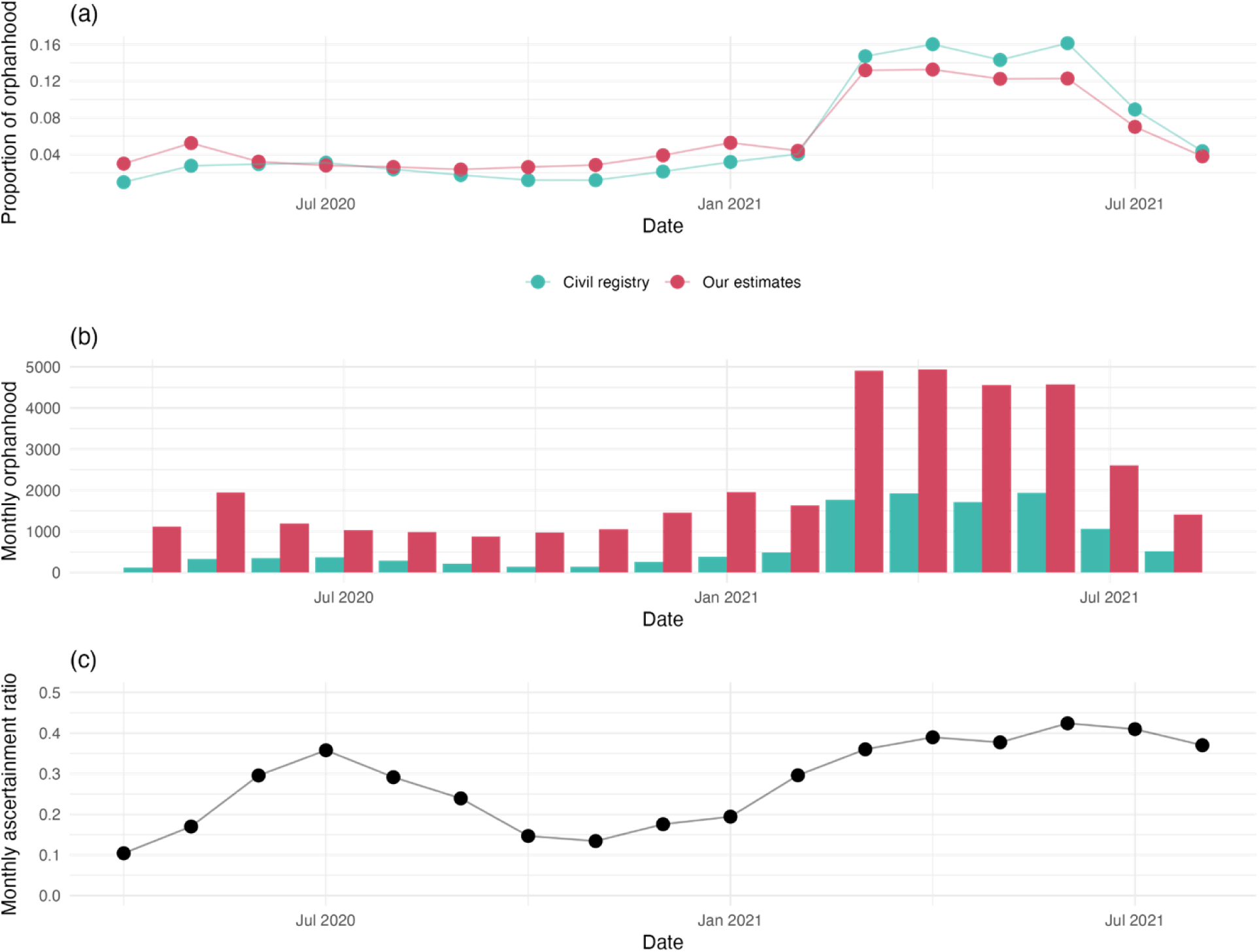
The proportion of COVID-19-associated orphanhood in 6-year-olds and under that occurred from April 2020 through August 2021 by month and source of data (panel a), monthly orphanhood (panel b), and monthly ascertainment ratio of civil registry data relative to our estimates (panel c).

However, the magnitude of our model estimates differed from the registry data. Using our monthly COVID-19-associated orphanhood methods and adjusting so the total matches the central estimates from the full model, we estimate 37,200 children aged 6 and under experienced orphanhood from 1 April 2020 through 31 August 2021. Dividing the 12,010 from the Arpen-Brasil dataset by 37,200 yields a civil registry ascertainment rate of 32.3%. We further estimate that, over this period, 27,100 children aged 6 and under lost a father and 10,100 children aged 6 and under lost a mother. The civil registry data estimates 8,079 children as having lost a father and 3,928 as having lost a mother, corresponding to ascertainment rates of 29.8% (for paternal orphanhood) and 38.9% (for maternal orphanhood). Ascertainment also varied by month, with lower values in early-2020 (10.4% in April 2020, the month with lowest ascertainment) and late-2020 (13.4% in November 2020), and higher values in mid-2020 (35.8% in July 2020) and 2021 (42.4% in June 2021, the month with the greatest ascertainment).

#### Campinas data

Manual review of death certificates in Campinas found that from 481 children, 434 children lost a mother or father, 1 lost a grandfather, and 46 lost an unknown caregiver from COVID-19. Of the 434 that lost a mother or father, 293 (67.5%) were listed as having lost a father, and 141 (32.5%) were listed as having lost a mother. These are similar proportions of paternal versus maternal orphans to our estimates and to those from Arpen-Brasil. These data also support our conclusions that orphanhood occurs more in older children (supplementary section 3.5).

Campinas, population 1.14m, accounts for 2.5% of São Paulo State and 2.4% of children aged 0 to 19 (“Panorama do Censo 2022” 2022). Multiplying our estimates of orphanhood in São Paulo State by 2.4% yields 543 paternal orphans, 233 maternal orphans, and 1 double orphan. This corresponds to an ascertainment rate of 54% for paternal orphans, 61% for maternal orphans, and 56% overall.

## Discussion

In Brazil in 2020-21, we estimated that 673,000 children (more than 1 in 100 total) experienced orphanhood, and a total of 1.3 million children experienced death of parents, co-residing grandparents, or older kin. Orphanhood and caregiver loss are a permanent loss, and although alternative care arrangements have been found to alleviate negative outcomes, provision and careful planning is needed in both the short and longer term. Loss of caregivers is a recognized ACE, during the pandemic deaths of caregivers were especially severe affecting millions of children and youth. During that time, social support and bereavement processes were disrupted. School closures, health care diversion, and social isolation severely interrupted essential pathways for care, support and attention. Cultural mourning and bereavement experiences were also restricted by pphysical distancing measures as was the ability to seek help. COVID-19 specifically caused a quarter of parental deaths, leaving in its wake 149,000 children experiencing orphanhood in the first two years of the pandemic, and 284,000 total children bereft of a parent or co-residing grandparent and other older kin. These bereavements will endure.

Beyond COVID-19, all-cause orphanhood as a major public health threat is escalating in the poly-crisis era, thus emerging as a new priority. In the U.S., for example, recent reports suggest that over 4 percent of all children had experienced prevalent orphanhood over the two decades spanning 2000-2021, with drug overdose figuring as the major cause of orphanhood in 2021 – even exceeding COVID-19-linked orphanhood (Villaveces et al. 2025). In Brazil, further insights into potentially important causes of orphanhood beyond COVID-19 may be gleaned from age- and sex-specific mortality data: for example, given that interpersonal violence constitutes the top cause of death for women aged 30-34 in Brazil (WHO 2024), intimate partner violence may be an important cause of orphanhood for children bereft of mothers in this age range. Preliminary evidence for the African continent suggests over 10% of all children have experienced death of a mother or father (N’konzi et al. 2023). Survey data for Colombia, South America, also suggest a large orphanhood burden, with more than 12% of 18-24 year olds surveyed in 2018 having experienced orphanhood (Ministry of Health and Social Protection 2018).

In planning for future epidemics, it is important to note that COVID-19-associated causes were a large contributing factor to all-cause orphanhood and caregiver loss among children in Brazil. Our state specific estimates highlight substantial differences with the most affected state (Mato Grosso, 4.4 children per 1000 estimated to have lost one or both parents) having an estimated rate of orphanhood 3.1 times that of the least affected state (Pará, 1.4 per 1000). Brazil, the most populous and largest country in Latin America, is also socio-economically (e.g. housing and employment) and geographically (e.g. access to services) one of the most unequal (International Monetary Fund 2019). Prior studies looking at COVID-19 mortality have shown that socio-economic vulnerabilities were major drivers in mortality outcomes during the period of the pandemic (Rocha et al. 2021), beyond population age structure and prevalence of chronic diseases. Our findings from a geographic perspective are consistent with the findings of that prior research and highlight similar areas where socio-economic vulnerabilities are more salient, primarily in the Northern regions (Rocha et al. 2021; Pinho Neto et al. 2024). Our research further expands prior findings in that it addresses all-cause mortality among caregivers and its differential impact on orphanhood.

Many causes of orphanhood beyond COVID-19 are important to address from a prevention point of view. While the effects of that pandemic have largely subsided, other causes are ongoing. One such cause is violence in its multiple expressions which affects mostly younger populations many of which are likely to have younger children in their families. In its most lethal manifestation, Brazil ranks first in terms of counts of homicide deaths globally and has rates over 20 per 100,000 population in the American continent (“Chapter 2: Homicide Trends and Patterns” 2023). The Americas report the highest homicide rates per 100,000 population globally, mainly affecting younger males (ages 15-29 years-old) (“Chapter 2: Homicide Trends and Patterns” 2023). While in recent years, national trends show reductions in homicide, subnationally, trends are very different at State levels (“Global Study on Homicide” 2023).

While attention has been drawn to COVID-19 orphans in research publications (Hillis, Unwin, Chen, et al. 2021; Unwin et al. 2022) and policy briefs (Hillis, Unwin, Cluver, et al. 2021), especially in Brazil (CDC 2022), they have to date been based on modelling approaches, combining epidemiological, demographic, and statistical methods. While these methods are especially valuable in data poor settings, wherever possible these approaches should be augmented by analyses of directly-observed data, such as administrative or survey data. With the notable exception of Brazil, these data simply did not exist in the early years of the pandemic (and we are unaware of any comprehensive efforts subsequently.) The situation in Brazil thus presented a unique opportunity to validate the modeling methods and to characterize the degree to which administrative data undercounts orphans.

When comparing the fraction of maternal and paternal orphans in the Arpen-Brasil Civil Registry dataset and the Campinas dataset to our modeled estimates, there was a very close match. The same was true of a temporal match between our modeled estimates and the Arpen-Brasil dataset. However, there was a stark difference in the magnitude of the number of children affected. Arpen-Brasil’s numbers were about one third as high as our estimates, and the Campinas numbers about one half, both of which are to be expected with administrative data. There are two reasons why administrative data would provide an undercount of COVID-19 orphans. First, COVID-19 was itself undercounted in death records in the context of the unfolding pandemic in 2020-21 (Msemburi et al. 2023). Secondly, both the Arpen-Brasil and Campinas datasets that we used relied on death certificates. While Brazil is unique in the world in that death certificates record information on child dependents of the deceased (Flaxman et al. 2023), this field is not digitally available and requires local work manually checking each certificate, and we would never expect this type of administrative data collection to yield complete data. Similarly, the Arpen-Brasil dataset relied on a matching process carried out by individual Civil Registry offices across Brazil between death and birth certificates (a process which was on the whole only possible for children aged 0 to 6 due to recent changes in how identification numbers are now assigned at birth.) This type of matching with governmental records is never perfect, and birth certificates may fail to list fathers (Caulfield 2012). We also hypothesize that our modeled approaches will likely undercount the true number of orphans, which is why we consider them to be minimum estimates of children affected. For example, poorer parents are likely to have more children and thus more exposure to infectious diseases, increasing their risk of mortality.

Our methods also have limitations. Various simplifying probabilistic assumptions were made, such as the independence between the probability that both parents die, when in reality, co-residing family members share many exposures such as to infectious pathogens or environmental hazards. The independence assumption likely results in us undercounting double orphanhood, and thus slightly overcounting total orphanhood. We also assume that fertility and mortality are independent. If parents with more children were more likely to die, then we will underestimate the true rate of orphanhood (and vice-versa). As with any statistical analysis, we are also limited by the data that were collected. While the PNS survey (used for male fertility and household structure estimates) features a large sample size, there are still high levels of uncertainty when estimating quantities at the state-level or by the age-of-child. Multiple approximations were required when using other data sources also, such as the interpolation of population count data prior to 2010, and the application of point estimates of live births underreporting from a single year to multiple years of births data. These approximations are small and are expected to cause only small bias, but the extent of this bias is again difficult to quantify.

The COVID-19 pandemic serves as an example of government reaction and preparedness for a crisis. The sharp, rapid, and comprehensive response needed for children can only begin to be effective if it is based on accurate, targeted, and available data to guide the response. This study has important policy implications for governments who continue to struggle with how to effectively design and deliver social assistance to orphans. The very first step in policy is the accurate identification of the target population in need which will focus the response adequately. Official death certificates are effective in helping to identify orphans, but administrative data alone are insufficient. In Brazil in particular, the evidence presented in this study suggests that there are sizable shares of orphans in the groups of children 7 years and older, including adolescents, who are wholly missing from the nationwide administrative data. As we noted earlier, despite the magnitude of the number of caregivers lost to COVID-19, a national program to monitor and provide a broad array of services to orphans does not exist in Brazil. Most of the legislation introduced in 2021 is still under discussion in the Senate. This provision may serve as a global blueprint for child crisis provision. Addressing the limitations of information systems aimed at identifying and monitoring these children at risk may increase the accuracy of estimates and better inform public health responses. Urgent attention is warranted to refine these systems so that the response to the next crisis meets the needs of so many children who currently fall under the radar.

## Data availability

All code and data (where we are able to provide directly) required to reproduce these results are available at https://github.com/MLGlobalHealth/BrazilOrphanhood. Where we are unable to provide original data, instructions are provided in this repository on how to obtain the data.

## Ethics

The study used only open available data. Death certificates in Brazil are public documents (federal law n. 6015 issued on December 31, 1973) and were obtained and reviewed for Campinas by co-author Andrea Santos Souza. Administrative data on COVID-19 orphans were shared by the Associação Nacional dos Registradores de Pessoas Naturais (Arpen-Brasil).

## Acknowledgments

N.S. acknowledges support from the Oxford-Radcliffe Scholarship from University College, Oxford, the EPSRC CDT in Modern Statistics and Statistical Machine Learning (Imperial College London and University of Oxford), A. Maslov for studentship support, EPSRC EP/V002910/2, and US Centers for Disease Control and Prevention and World Health Organization, “Building Global Public Health Capacity to Link Real-Time Modelling Data on COVID-19-associated Orphanhood and Caregiver Deaths to Inform Prevention, Preparedness and Protection from COVID-19 consequences,” 2023 (CDC/WHO grant). H.T.J.U. acknowledges funding from the Moderna Charity Foundation. A.B. acknowledges funding from EPSRC EP/X038440/1. E.S. acknowledges funding from EPSRC EP/V002910/2 and support from the AI2050 program at Schmidt Sciences (Grant [G-22-64476]). A.V.R.A and O.R. acknowledge funding from the Moderna Charity Foundation. L.C. acknowledges funding from the European Research Council (#771468). L.B. acknowledges CNPq (Conselho Nacional de Desenvolvimento Científico e Tecnológico (CNPq)), Grant (312475/2022-5) and Fapesp (Fundação de Amparo à Pesquisa do Estado de São Paulo (Fapesp)), Grant (2021/08772-9). S.F. acknowledges funding from CDC/WHO grant and EPSRC EP/V002910/2.

We thank Jeff Imai-Eaton for helpful discussions about methods for analyzing survey data.

## References

Belsey, Mark A., and Lorraine Sherr. 2011. “The Definition of True Orphan Prevalence: Trends, Contexts and Implications for Policies and Programmes.” Vulnerable Children and Youth Studies 6 (3). Taylor & Francis: 185–200.

Brizzi, Andrea, Charles Whittaker, Luciana M. S. Servo, Iwona Hawryluk, Carlos A. Prete Jr, William M. de Souza, Renato S. Aguiar, et al. 2022. “Spatial and Temporal Fluctuations in COVID-19 Fatality Rates in Brazilian Hospitals.” Nature Medicine 28 (7): 1476–1485.

Buchanan, Ann, and Anna Rotkirch. 2018. “Twenty-First Century Grandparents: Global Perspectives on Changing Roles and Consequences.” Contemporary Social Science 13 (2). Informa UK Limited: 131–144.

Castro, Marcia C., Sun Kim, Lorena Barberia, Ana Freitas Ribeiro, Susie Gurzenda, Karina Braga Ribeiro, Erin Abbott, Jeffrey Blossom, Beatriz Rache, and Burton H. Singer. 2021. “Spatiotemporal Pattern of COVID-19 Spread in Brazil.” Science 372 (6544): 821–826.

Caulfield, Sueann. 2012. “The Right to a Father’s Name: A Historical Perspective on State Efforts to Combat the Stigma of Illegitimate Birth in Brazil.” Law and History Review 30 (1). Cambridge University Press (CUP): 1–36.

CDC. 2022. “Global Orphanhood Associated with COVID-19.” https://archive.cdc.gov/www_cdc_gov/globalhealth/covid-19/orphanhood/index.html.

“Chapter 2: Homicide Trends and Patterns.” 2023. In Global Study on Homicide 2023. Vienna: United Nations Office on Drugs and Crime.

Cluver, Lucie, Jeffrey W. Imai-Eaton, Lorraine Sherr, Mary Mahy, and Seth Flaxman. 2023. “Reauthorise PEPFAR to Prevent Death, Orphanhood, and Suffering for Millions of Children.” The Lancet 402 (10404): 769–770.

“Covid já deixou órfãs ao menos 12 mil crianças de até 6 anos, indicam cartórios.” 2021. *ISTOÉ*. https://istoe.com.br/covid-ja-deixou-orfas-ao-menos-12-mil-criancas-de-ate-6-anos-indicam-cartorios/.

De Kerf, Joseph L. F. 1975. “The Interpolation Method of Sprague-Karup.” Journal of Computational and Applied Mathematics 1 (2): 101–110.

Evans, David K., and Anna Popova. 2015. Orphans and Ebola: Estimating the Secondary Impact of a Public Health Crisis. The World Bank.

Faria, Nuno R., Thomas A. Mellan, Charles Whittaker, Ingra M. Claro, Darlan da S. Candido, Swapnil Mishra, Myuki A. E. Crispim, et al. 2021. “Genomics and Epidemiology of the P.1 SARS-CoV-2 Lineage in Manaus, Brazil.” Science 372 (6544): 815–821.

Flaxman, Seth, Lackson Kasonka, Lucie Cluver, Andrea Santos Souza, Charles A. Nelson 3rd, Alexandra Blenkinsop, H. Juliette T. Unwin, and Susan Hillis. 2023. “List Child Dependents on Death Certificates.” Science 380 (6644): 467.

Gangavarapu, Karthik, Alaa Abdel Latif, Julia L. Mullen, Manar Alkuzweny, Emory Hufbauer, Ginger Tsueng, Emily Haag, et al. 2023. “Outbreak.Info Genomic Reports: Scalable and Dynamic Surveillance of SARS-CoV-2 Variants and Mutations.” Nature Methods 20 (4): 512–522.

“Global Study on Homicide.” 2023. *United Nations : Office on Drugs and Crime*. https://www.unodc.org/unodc/en/data-and-analysis/global-study-on-homicide.html.

Goldman, Philip S., Marian J. Bakermans-Kranenburg, Beth Bradford, Alex Christopoulos, Patricia Lim Ah Ken, Christopher Cuthbert, Robbie Duchinsky, et al. 2020. “Institutionalisation and Deinstitutionalisation of Children 2: Policy and Practice Recommendations for Global, National, and Local Actors.” The Lancet. Child & Adolescent Health 4 (8). Elsevier BV: 606–633.

Hillis, Susan D., H. Juliette T. Unwin, Yu Chen, Lucie Cluver, Lorraine Sherr, Philip S. Goldman, Oliver Ratmann, et al. 2021. “Global Minimum Estimates of Children Affected by COVID-19-Associated Orphanhood and Deaths of Caregivers: A Modelling Study.” The Lancet 398 (10298): 391–402.

Hillis, Susan D., Juliette Unwin, Lucie Cluver, Lorraine Sherr, Philip Goldman, Laura Rawlings, Gretchen Bachman, et al. 2021. “Children: The Hidden Pandemic 2021: A Joint Report of COVID-19-Associated Orphanhood and a Strategy for Action.” https://stacks.cdc.gov/view/cdc/108199.

IBGE. 2018. “Population Projection.” https://www.ibge.gov.br/en/statistics/social/population/18176-population-projection.html?edicao=21933.

IBGE. 2022. “Estudo complementar à aplicação da técnica de captura-recaptura : estimativas desagregadas dos totais de nascidos vivos e óbitos : 2020 / IBGE, Coordenação de População e Indicadores Sociais.” https://biblioteca.ibge.gov.br/index.php/biblioteca-catalogo?view=detalhes&id=2101978.

International Monetary Fund. 2019. Brazil: Boom, Bust, and the Road to Recovery. Edited by Antonio Spilimbergo and Krishna Srinivasan. London, England: International Monetary Fund.

Kessler, Ronald C., Katie A. McLaughlin, Jennifer Greif Green, Michael J. Gruber, Nancy A. Sampson, Alan M. Zaslavsky, Sergio Aguilar-Gaxiola, et al. 2010. “Childhood Adversities and Adult Psychopathology in the WHO World Mental Health Surveys.” The British Journal of Psychiatry: The Journal of Mental Science 197 (5): 378–385.

Kidman, Rachel, and Tia Palermo. 2016. “The Relationship between Parental Presence and Child Sexual Violence: Evidence from Thirteen Countries in Sub-Saharan Africa.” Child Abuse & Neglect 51 (January): 172–180.

“Lei Ordinária 16200 2022 de Campinas SP.” 2022. https://leismunicipais.com.br/a/sp/c/campinas/lei-ordinaria/2022/1620/16200/lei-ordinaria-n-16200-2022-institui-plano-de-acao-destinado-as-criancas-e-aos-adolescentes-em-situacao-de-orfandade-causada-pela-covid-19-no-municipio-de-campinas.

Maxmeio. 2021. “Governadores do Nordeste lançam programa “Nordeste Acolhe”, que prevê beneficio de R$500 aos órfãos da Covid-19.” *Consórcio Nordeste*. https://www.consorcionordeste.gov.br/noticia/governadores-do-nordeste-lancam-programa-nordeste-acolhe-que-preve-beneficio-de-r500-aos-orfaos-da-covid-19.

Ministry of Health and Social Protection. 2018. *Colombia Violence Against Children and Youth Survey*. Government of Colombia. https://cdn.togetherforgirls.org/assets/files/Colombia-VACS-Report.pdf.

Msemburi, William, Ariel Karlinsky, Victoria Knutson, Serge Aleshin-Guendel, Somnath Chatterji, and Jon Wakefield. 2023. “The WHO Estimates of Excess Mortality Associated with the COVID-19 Pandemic.” Nature 613 (7942): 130–137.

N’konzi, J. P. N., L. Cluver, S. Hillis, S. Flaxman, and H. J. T. Unwin. 2023. “All-Cause Orphanhood Prevalence & Incidence for Children-Inclusive Policy Planning in Africa.” In 22nd International Conference on AIDS & STIs in Africa. Poster presentation.

OPENDATASUS. 2022. “Sistema de Informação sobre Mortalidade – SIM.” https://opendatasus.saude.gov.br/dataset/sim.

“Panorama do Censo 2022.” 2022. *Panorama do Censo* 2022. https://censo2022.ibge.gov.br/panorama/?utm_source=ibge&utm_medium=home&utm_campaign=portal.

Pinho Neto, Valdemar, Cecilia Machado, Felipe Lima, Soraya Roman, and Gilson Dutra. 2024. “Inequalities in the Geographic Access to Delivery Services in Brazil.” BMC Health Services Research 24 (1): 1598.

“PL 2291/2021 - Senado Federal.” 2023. https://www25.senado.leg.br/web/atividade/materias/-/materia/148882.

Ponmattam, Jamie, Andrew Stokes, Lucas Carvalho, Cassio Turra, Sun Kim, and Marcia Castro. 2024. “Covid-19 and Excess Mortality in Brazil: Subnational Estimates for 2020 and 2021.” *In Preparation*.

“Portal da Câmara dos Deputados.” 2021a. https://www.camara.leg.br/proposicoesWeb/fichadetramitacao/?idProposicao=2278389.

“Portal da Câmara dos Deputados.” 2021b. https://www.camara.leg.br/proposicoesWeb/fichadetramitacao/?idProposicao=2283096.

“Portal da Câmara dos Deputados.” 2023. https://www.camara.leg.br/proposicoesWeb/fichadetramitacao?idProposicao=2346821.

Rocha, Rudi, Rifat Atun, Adriano Massuda, Beatriz Rache, Paula Spinola, Letícia Nunes, Miguel Lago, and Marcia C. Castro. 2021. “Effect of Socioeconomic Inequalities and Vulnerabilities on Health-System Preparedness and Response to COVID-19 in Brazil: A Comprehensive Analysis.” The Lancet. Global Health 9 (6): e782–e792.

Sherr, Lorraine, Rebecca Varrall, Joanne Mueller, JLICA Workgroup 1 Members, Linda Richter, Angela Wakhweya, Michele Adato, et al. 2008. “A Systematic Review on the Meaning of the Concept ‘AIDS Orphan’: Confusion over Definitions and Implications for Care.” AIDS Care 20 (5): 527–536.

“Sistema de Informação Sobre Nascidos Vivos – Sinasc - OPENDATASUS.” 2023. https://opendatasus.saude.gov.br/en/dataset/sistema-de-informacao-sobre-nascidos-vivos-sinasc.

“Survey: Analysis of Complex Survey Samples.” 2023. *Comprehensive R Archive Network (CRAN)*. https://cran.r-project.org/web/packages/survey/.

Thomas, Tina, Mei Tan, Yusra Ahmed, and Elena L. Grigorenko. 2020. “A Systematic Review and Meta-Analysis of Interventions for Orphans and Vulnerable Children Affected by HIV/AIDS Worldwide.” Annals of Behavioral Medicine: A Publication of the Society of Behavioral Medicine 54 (11): 853–866.

Timæus, I. 2021. “The Own-Children Method of Fertility Estimation: The Devil Is in the Detail.” Demographic Research 45 (September): 825–840.

Unwin, H. Juliette T., Susan Hillis, Lucie Cluver, Seth Flaxman, Philip S. Goldman, Alexander Butchart, Gretchen Bachman, et al. 2022. “Global, Regional, and National Minimum Estimates of Children Affected by COVID-19-Associated Orphanhood and Caregiver Death, by Age and Family Circumstance up to Oct 31, 2021: An Updated Modelling Study.” The Lancet. Child & Adolescent Health 6 (4): 249–259.

Villaveces, Andrés, Yu Chen, Sydney Tucker, Alexandra Blenkinsop, Lucie Cluver, Lorraine Sherr, Jan L. Losby, et al. 2025. “Orphanhood and Caregiver Death among Children in the United States by All-Cause Mortality, 2000-2021.” Nature Medicine, January. Springer Science and Business Media LLC. doi:10.1038/s41591-024-03343-6.

WHO. 2024. “Global Health Estimates.” https://www.who.int/data/global-health-estimates.

